# Purifying selection determines the short-term time dependency of evolutionary rates in SARS-CoV-2 and pH1N1 influenza

**DOI:** 10.1101/2021.07.27.21261148

**Authors:** Mahan Ghafari, Louis du Plessis, Jayna Raghwani, Samir Bhatt, Bo Xu, Oliver G Pybus, Aris Katzourakis

## Abstract

High throughput sequencing enables rapid genome sequencing during infectious disease outbreaks and provides an opportunity to quantify the evolutionary dynamics of pathogens in near real-time. One difficulty of undertaking evolutionary analyses over short timescales is the dependency of the inferred evolutionary parameters on the timespan of observation. Here, we characterise the molecular evolutionary dynamics of SARS-CoV-2 and 2009 pandemic H1N1 (pH1N1) influenza during the first 12 months of their respective pandemics. We use Bayesian phylogenetic methods to estimate the dates of emergence, evolutionary rates, and growth rates of SARS-CoV-2 and pH1N1 over time and investigate how varying sampling window and dataset sizes affects the accuracy of parameter estimation. We further use a generalised McDonald-Kreitman test to estimate the number of segregating non-neutral sites over time. We find that the inferred evolutionary parameters for both pandemics are time-dependent, and that the inferred rates of SARS-CoV-2 and pH1N1 decline by ∼50% and ∼100%, respectively, over the course of one year. After at least 4 months since the start of sequence sampling, inferred growth rates and emergence dates remain relatively stable and can be inferred reliably using a logistic growth coalescent model. We show that the time-dependency of the mean substitution rate is due to elevated substitution rates at terminal branches which are 2-4 times higher than those of internal branches for both viruses. The elevated rate at terminal branches is strongly correlated with an increasing number of segregating non-neutral sites, demonstrating the role of purifying selection in generating the time-dependency of evolutionary parameters during pandemics.

## 1. Introduction

Rapid whole-genome sequencing has become part of pathogen surveillance systems and is important to both infection control and enables a detailed investigation of the epidemiological and evolutionary characteristics of pathogens. Quantifying infectious disease evolution enables the inference of parameters such as times of origin, epidemic growth rates, and evolutionary rates^1-3^.

One of the perils of making such inferences over short time periods (i.e., a few months or years) is that the inferred parameters of interest may vary over the timespan of observation^4^. This nonstationarity can render findings that are confusing or conflicting such that, for instance, an estimated substitution rate over one time window is not transferable to the analysis of another^5^. While several factors can lead to the misestimation of substitution rates (such as mis-identification of coalescent population models, changes in natural selection, differing replication rates in various host reservoirs, or sequencing errors), misspecification of clock models and saturation of nucleotide changes can result in rate underestimation^6-9^.

Another major obstacle is that during an ongoing outbreak the level of sequence diversity may be so low that the amount of accrued evolutionary change is not sufficient to make informative phylogenetic inference possible^10^. Several statistical tests have been developed to ensure enough temporal signal is present in a set of temporally sampled genome sequences to reliably infer evolutionary parameters^11,12^.

More specifically, there is extensive evidence that purifying selection in viruses can result in varied estimates for rates of substitution and evolutionary rate ratio, dN/dS, across different timescales^13-15^. It is likely that a greater proportion of polymorphisms observed among genomic sequences sampled early in an epidemic are segregating deleterious mutations, which will only persist for a limited time before being eliminated by purifying selection^16^. The duration of this time-dependent effect may be prolonged due to incomplete purifying selection in populations with very large effective population sizes^17^.

While evidence of strong purifying selection has mostly been reported at the species level and over very long timescales (i.e., thousands to millions of years), using inference methods based on the dN/dS ratio^18-20^, there have been few studies at the intra-population level and over short timescales, mainly because there is often no opportunity to collect sufficiently large number of samples through time to track low-frequency variants. This has changed recently due to the rapid sharing of pathogen genome data on platforms such as GISAID and GenBank. Furthermore, using standard phylogenetic methods to compute the dN/dS ratio for conspecific sequences sampled from a single population over short timescales may be inappropriate as the differences between sequences over such timescales represent segregating polymorphisms as opposed to fixed substitutions along independent lineages. The former has been shown to produce very different estimates of the dN/dS ratio over time^21,22^.

In this study, we aim to quantify and identify the source of time-dependency of virus substitution rate estimates over short time periods and characterise estimates of the molecular clock rate, time of origin, growth rate, and number of non-neutral sites for different datasets that represent different timescales of genomic observation. We show that time-dependency in estimates of the mean substitution rate is dominated by elevated rates at terminal branches, whereas the estimated rate of substitution at internal branches does not exhibit a time-dependent decay. We then use a generalised McDonald-Kreitman test based on nucleotide site frequencies that allows purifying selection to be quantified over short timescales and demonstrate that there is a strong correlation between the elevated rates at terminal branches and the high number of low frequency non-neutral sites in both SARS-CoV-2 and pH1N1 genomes.

## 2. Results

We use Bayesian phylogenetic methods implemented in BEAST v.1.10 to estimate the molecular clock rate, times of origins, and growth rates of SARS-CoV-2 and pH1N1 pandemics. The dataset varied in size and temporal sampling range and were created to reflect the way in which real datasets accumulate in size and diversity during the course of an epidemic. Specifically, we aggregate all available samples up to each respective month and infer the parameters of interest (**Figure 1; Figure S1**). We first compare the results for SARS-CoV-2 using two coalescent growth priors: exponential growth and logistic growth. We find that, except for the month of January, the logistic growth coalescent tree prior is a better fit to the data (**Table 1**). As reported in previous studies, we find that there is not enough temporal signal in the data to reliably infer the evolutionary parameters during the first two months of the SARS-CoV-2 pandemic^10^. This results in the underestimation of substitution rates as well as high statistical uncertainty for the parameter estimates. The lack of temporal signal in the SARS-CoV-2 samples is also evident from the number of new singletons (i.e., single nucleotide variations in the dataset compared to the ancestral sequence) added the dataset per month during the first 2-3 months (**Figure S2**). On the other hand, for pH1N1, the molecular clock rate is up to 2 times higher for the first three months than for the following months (**Figure 1 A, B**). While the inferred substitution rate of SARS-CoV-2 tends to decrease as we increase the timespan of measurement, the rate for pH1N1 does not change considerably after the first 3-4 months of measurement, in agreement with previous findings^4^.

**Table 1:**
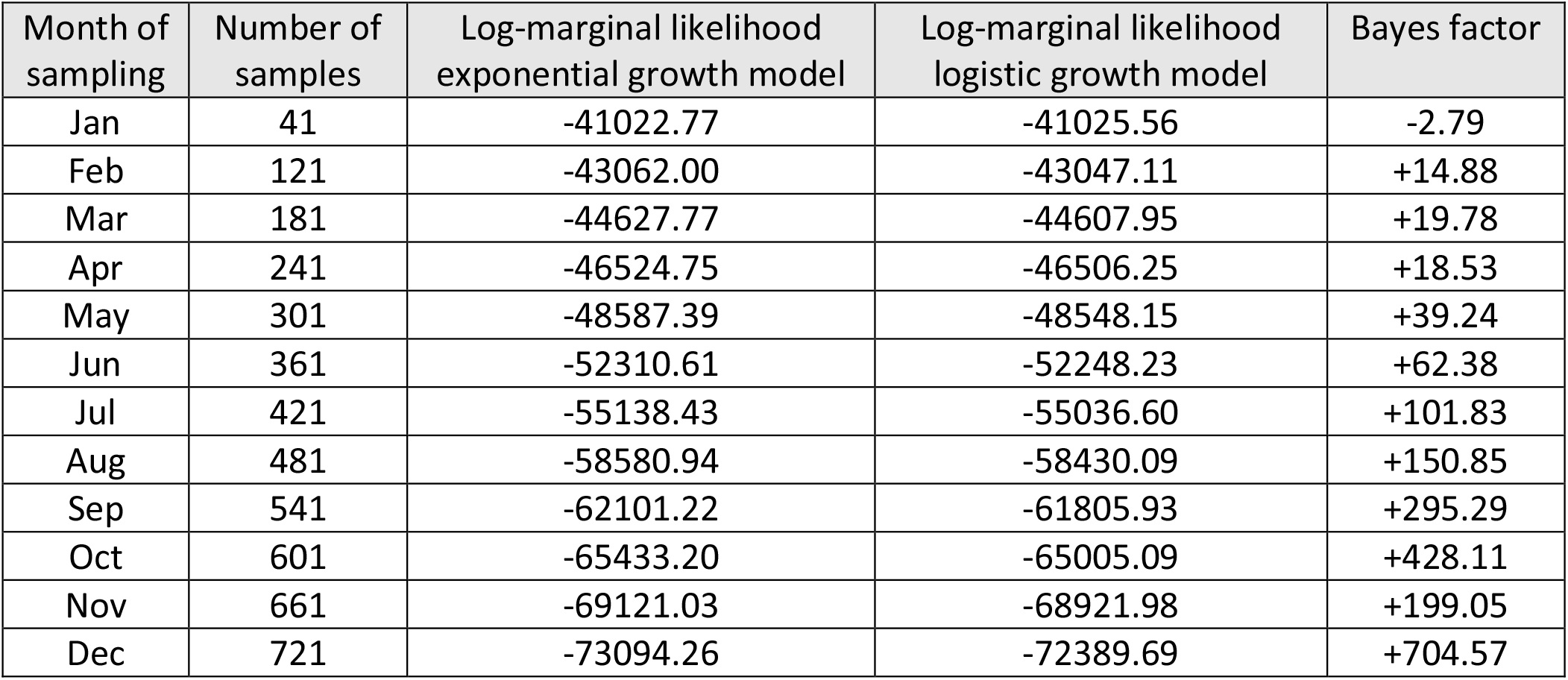
Log-marginal likelihoods of exponential and logistic growth models with increasing temporal ranges of sampling dates. Taking exponential growth as the null model, we select the logistic growth model for any dataset with a positive Bayes factor.

**Figure 1:**
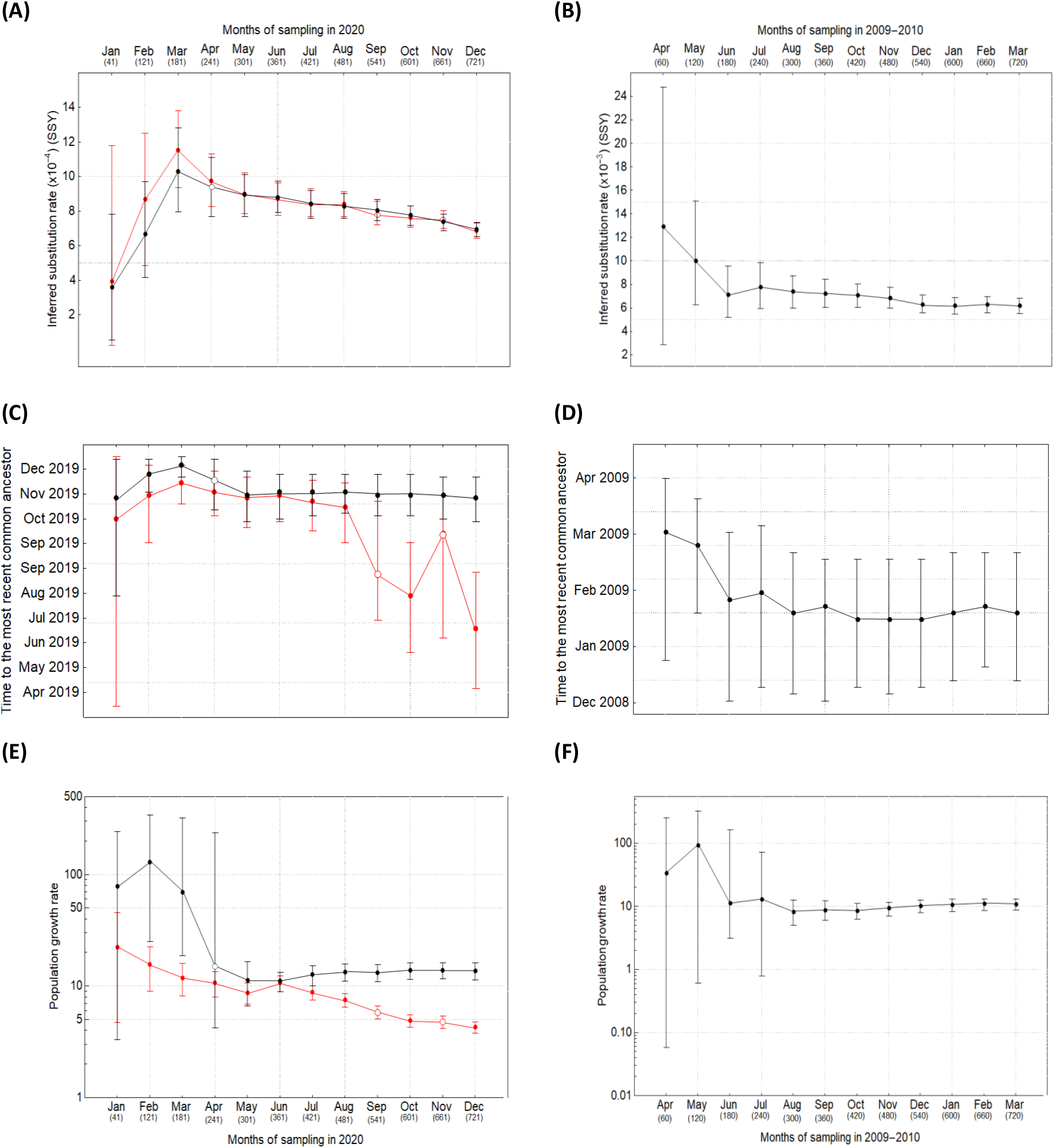
(**A, B**) Inferred rates, (**C, D**) times of origin, and (**E, F**) growth rates of SARS-CoV-2 (left column) and pH1N1 influenza (right column) using an exponential (black) and a logistic (red) growth model. Open circles represent nonconvergence for at least one parameter in the Bayesian analysis. Note that y-axes are not on the same scale for SARS-CoV-2 and pH1N1.

Both the inferred times of origin and growth rates of SARS-CoV-2 and pH1N1 in early months have wide confidence intervals due to the high uncertainty associated with small sample sizes and narrow sampling windows. The precision of these two inferred parameters increases with the addition of data from later months and remain roughly consistent after the first 4-5 months of measurement (**Figure 1 C-F**). Using the logistic growth model, we find the estimated times of origin for SARS-CoV-2 and pH1N1 to be 2019-10-28 (95% HPD: 2019-09-30, 2019-11-24) and 2009-01-18 (95% HPD: 2008-12-14, 2009-02-22), respectively, which is also in agreement with previous studies^2,3,23^. However, under the exponential growth model, the inferred time of origin of SARS-CoV-2 samples significantly diverge from the expectation after 6 months of sampling and yield unreliable estimates (**Figure 1 C, D**). This is likely because after the first few months, the growth rate declines and the population dynamics deviates from the exponential growth model (**Figure 1 E, F**). This, in turn, results in the underestimation of growth rate of SARS-CoV-2 and inferring an older time of origin.

Purifying selection has been often cited as one of the main evolutionary processes contributing to the elevation of estimated molecular evolutionary rates over short timescales^23^. The argument is that low-frequency deleterious mutations can segregate in a population for some time before being purged because of purifying selection. Therefore, the proportion of all changes that are deleterious is high when the phylogenetic tree is short, and lower when the tree is longer as it takes more time for shared neutral or advantageous changes to accrue with respect to the ancestral state. To evaluate this hypothesis, we investigate the correlation between the number of non-neutral polymorphisms and estimated substitution rates over time. Our results show that there is a higher proportion of segregating mutations at low frequencies (<15%) during the first few months of observation, in both the ORF1b gene of SARS-CoV-2 and the HA gene of pH1N1 (**Figure 2 A, B**). In particular, the proportion of low frequency non-neutral sites in ORF1ab gene was higher during the first 4-5 months of observation and had a roughly fourfold drop during that period compared to a twofold drop in HA gene. After the first 5 months, the proportion of low frequency non-neutral sites remains roughly constant at around 15% in ORF1ab, while it shows an uptick from 15% to 17% in HA towards the beginning of 2010. The slight increase in the proportion of non-neutral sites in HA during this period is also in agreement with a rise in the relative genetic diversity of pH1N1 around the world in late 2009/early 2010 which may also have resulted in an increase in the number of segregating deleterious mutations in the population^24^. We also find that the number of low frequency replacement sites is always greater than silent sites in ORF1ab while, for HA, silent sites are in majority in most months (**Figure 2 C, D**). The total number of non-neutral sites in ORF1ab is higher because it is a much longer gene (∼21,000 nt) compared to HA (∼1,800 nt).

**Figure 2:**
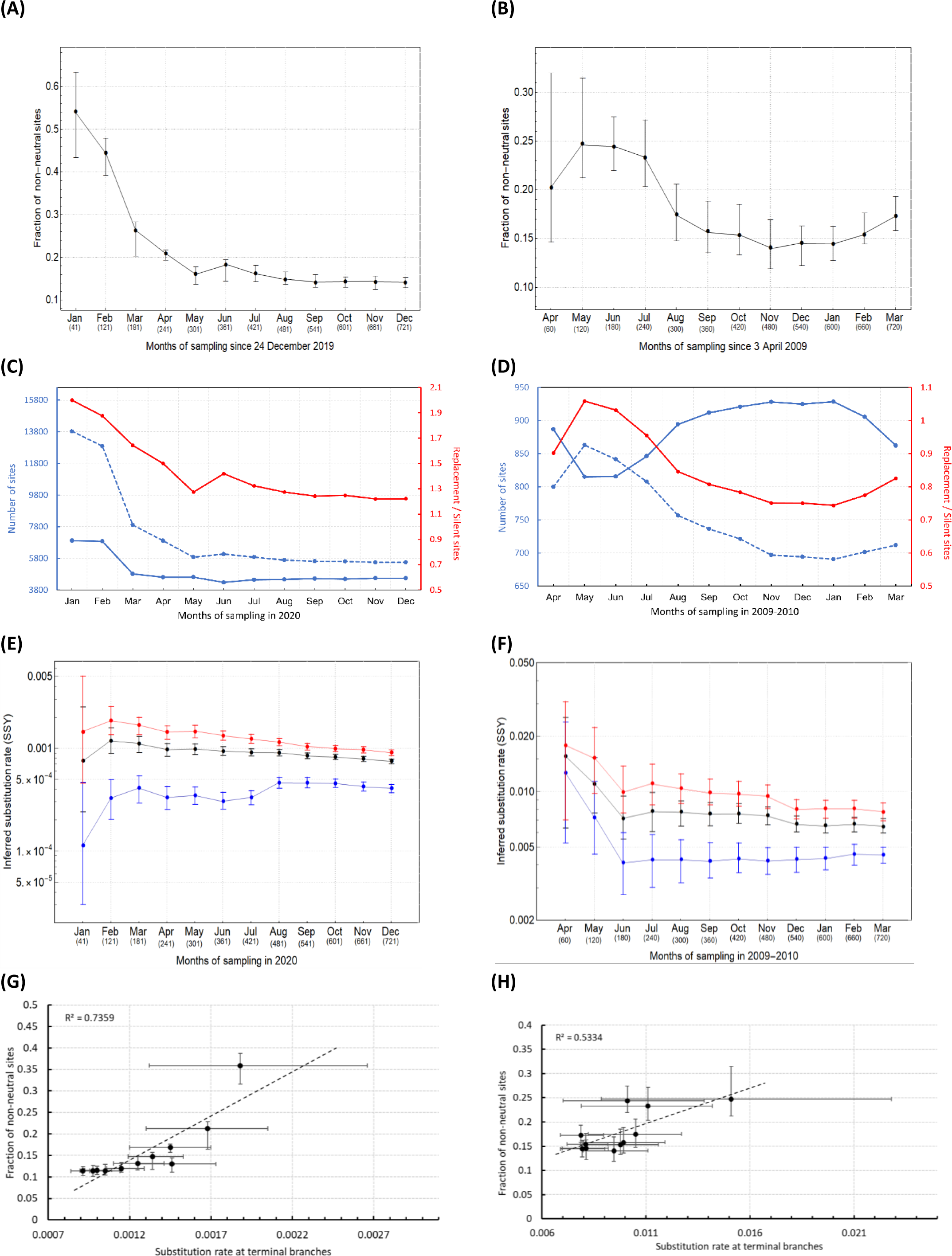
**(A, B)** Number of non-neutral sites over time for SARS-CoV-2 (left column) and pH1N1 (right column). **(C, D)** Number of replacement (dashed blue line), silent (solid blue line) sites, and their ratio (red line) over time. **(E, F)** Mean clock rate (black), and the rates at the terminal (red) and internal (blue) branches. The MCMC chains for the first month of sampling SARS-CoV-2 and the first three months of pH1N1 do not converge using the logistic growth coalescent model. Instead, the exponential growth coalescent model was used. **(G, H)** Correlation coefficient between the substitution rate at terminal branches and number of non-neutral sites -- excluding the estimates from the first month of sampling due to inadequate temporal signal and significant uncertainty in the inferred parameters.

One of the impacts of purifying selection over short timescales is that the number of replacements on terminal branches should be higher than that on internal branches. To measure this effect during the SARS-CoV-2 and pH1N1 epidemics, we used a branch-specific two parameter molecular clock model in BEAST to infer separate rates of substitution for terminal and internal branches (see **Figure 2 E, F**). Our results show that for both viruses, the rate of substitution on terminal branches are 2-5 times higher than on internal branches. The difference is more dramatic in SARS-CoV-2 where the average ratio of substitution rates at terminal branches are 4 times higher than that of internal branches while it is only 2 times higher in HA during the first 12 month of observation. This also agrees with the observation of a higher proportion of non-neutral sites in SARS-CoV-2 samples compared to pH1N1 because there is going to be more deleterious mutations at terminal branches of SARS-CoV-2. Further, there is a clear decline in the estimated rate at terminal branches as the sampling window increases. In contrast, the estimated rate at internal branches gradually increases through time (with the exception of the first 2-3 months). In **Figure 2 G and H**, we show that there is an overall significant correlation between the rate of substitution at terminal branches and number of non-neutral sites in both SARS-Cov-2 (R^2^=0.74 and p < 0.001) and pH1N1 (R^2^=0.53 and p < 0.01) (**Figure 2 G and H**). By examining the site frequency spectrum for both datasets, we can also see that most of the variation comes from the low-frequency regime (<15%) with very limited variation in the mid-to high-frequency regimes (**Figure S2**).

Furthermore, in the pH1N1 dataset, we can see a dip in the inferred rate at terminal branches in December 2009 (**Figure 2 F**). There is also a sudden loss of low frequency genetic diversity in the dataset during the same period which is partially recovered in February and March (see the trajectory of newly added singletons and low frequency variants in **Figure S2 D, F**). On the other hand, the pattern of substitution rate at internal branches is non-monotonic over time. In particular, we see an increase in the rate at internal branches of SARS-CoV-2 from August onwards. Similarly, there is a steady rise in the rate at internal branches of pH1N1 from February onwards.

## 3. Discussion

We found a strong correlation between the over-abundance of deleterious mutations during the early stages of both the SARS-CoV-2 and pH1N1 pandemics which results in higher substitution rates at terminal branches relative to internal branches of inferred phylogenetic trees. Once there is enough temporal signal in the data to reliably estimate relevant epidemiological and evolutionary parameters, the mean substitution rates decline over the course of one year of rate measurement while the estimated time of origin and growth rate are more stable and remain roughly the same after the first 4-5 months of measurement. We found that this declining pattern in mean substitution rate is caused by the rate decay at terminal branches which is also strongly correlated with the declining proportion of low frequency non-neutral sites over time.

Our results provided further evidence that short-term rate estimates are subject to time-dependent rate effects largely due to transient polymorphisms^4,23^. In addition, our estimated mean substitution rates, times of origin, and growth rates agree with previous studies of SARS-CoV-2^3,10,25,26^ and pH1N1^2,4,23,27^. We found that while the time-dependent rate effect for pH1N1 is less pronounced after the first 3 months of sampling, the same effect in SARS-CoV-2 is more pronounced and continues even after 12 months of sampling. This may be the result of a much higher mean substitution rate in pH1N1 compared to SARS-CoV-2. We also found a higher proportion of low frequency non-neutral sites for SARS-CoV-2 dataset during the first 8 months of observations and a higher ratio of replacement to silent sites during the entire one year of observation compared to pH1N1.

By dividing the data into successively longer temporal intervals, we showed that the over-abundance of deleterious mutations at terminal branches are the main reason behind the gradual decay in mean substitution rate over time in SARS-CoV-2 and pH1N1. Further work can be done to quantify the long-term effect of purifying selection on the time-dependency of substitution rates.

## 4. Methods

We downloaded all SARS-CoV-2 sequences from GISAID and pH1N1 influenza sequences from GenBank and align them using MUSCLE v3.8.425^28^ -- a complete metadata table acknowledging the authors, originating and submitting laboratories of the SARS-CoV-2 sequence data is available in **Table S1**. For the SARS-CoV-2 dataset, we mask sites in the first and last 130 bp of the alignments and only include complete sequences that are more than 29,000 nt long with high coverage as determined by GISAID’s default search option (i.e., entries with <1% Ns and <0.05% unique amino acid substitutions). We specifically select pH1N1 sequences from GenBank for the full coding region of the hemagglutinin (HA) gene.

To investigate the effects of the temporal range of sampling dates on the accuracy of parameter estimation, we incrementally increase the size of each dataset by adding 60 genomes, chosen randomly, for every additional month of sampling. Thus, after 12 months of sampling since the first sequence was uploaded on GISAID and/or GenBank, we have 720 samples. We note that due to a lack of temporal signal in the early SARS-CoV-2 samples (i.e., most of the early samples were almost completely identical) and failure of the Markov Chain Monte Carlo (MCMC) chains for the phylogenetic analyses to converge, we only used 41 samples collected between 24 December 2019 to 31 January 2020 (labelled as ‘January sequences’ in our analysis) and took 20 additional samples during the next month to match with the 120 samples used for pH1N1 by the end of the second month of sampling (see **Figure S1**).

### 4.1 Phylogenetic analyses

We use BEAST v1.10^29^ for the Bayesian phylogenetic analysis of the entire dataset using an HKY+Γ substitution model with a Laplace prior (mean=0 and scale=100) on the coalescent growth rate, a Lognormal prior (mean=1 and stdev=2) on the coalescent population size, and a continuous time Markov chain prior on the evolutionary clock rate. For the first part of the analysis, we use a strict clock model and exponential and logistic growth coalescent demographic models of SARS-CoV-2 evolution and only the logistic growth model for the HA segment of pH1N1. To quantify the relative fit of the two coalescent models for SARS-CoV-2, we compute their log marginal likelihoods using the generalised stepping-stone sampling method and compare their Bayes factors^30,31^.

For each dataset, we perform MCMC runs for 100 million steps, sample trees every 10 thousand steps, and remove the first 10% of the steps as burin-in. We ensure that the effective sample size for every parameter of interest is >200 using Tracer v1.7^32^. For the second part of the analysis, we use a two-parameter molecular clock model with one clock rate for terminal branches and one for internal branches, using the same priors as before. We use an MCMC chain of length 50 million steps, sampling every 1000 steps and evaluate sampling of the parameters of interest using Tracer v1.7.

### 4.2 Estimating the site frequency and number of non-neutral sites

We use the adapt-a-rate package^33-35^, a generalised McDonald-Kreitman test, to estimate the number of non-neutral sites by assuming that deleterious mutations are mostly confined to the low frequency range (0%-15%), neutral mutations to the mid frequency range (15%-75%), and adaptive mutations to the high frequency range (75%-100%). We then estimate the site frequency spectra by comparing the main alignments to an ancestral sequence. For SARS-CoV-2, the ancestral sequence is the earliest sample collected from Wuhan, Wuhan/IPBCAMS-WH-01/2019, and for pH1N1 it is the earliest sample from Mexico, ACQ99614|A/Mexico/4108/2009. We use the ORF1ab gene of SARS-CoV-2 and HA gene of pH1N1 influenza for this analysis. We also carried out a similar analysis for the other genes of SARS-CoV-2, including the S gene. However, because of the limited genetic diversity present in the sequences and their relatively short size, our generalised McDonald-Kreitman test using adapt-a-rate is unable to estimate the number of non-neutral sites. The reason for choosing to analyse the HA gene of pH1N1 is primarily because of the availability of a large number of sequences from GenBank for the full coding region of HA which allows us to randomly sample 60 alignments per month from April 2009 to March 2010.

## Data Availability

All relevant data is available

https://github.com/mg878/twoclock_rate

https://github.com/jnarag/teaspoon

## Data and code availability

Datasets required to reproduce the analyses are available at https://github.com/mg878/twoclock_rate. The adapt-a-rate package is also available at https://github.com/jnarag/teaspoon.

## Acknowledgement

MG is funded by the Biotechnology and Biological Science Research Council (BBSRC), grant number BB/M011224/1.

## Supplementary Figures

**Figure S1:**
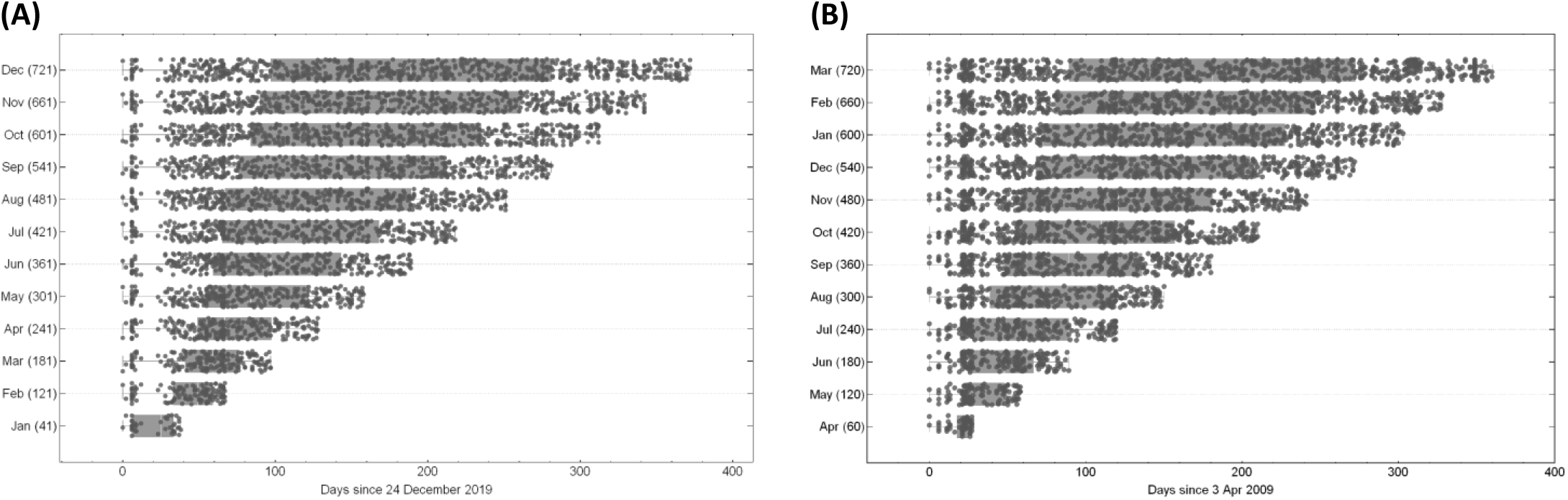
Sampling intervals and numbers of sequences in the different datasets used for SARS-CoV-2 and pH1N1 analyses. The sampling interval is measured from the day when the first genome was collected.

**Figure S2:**
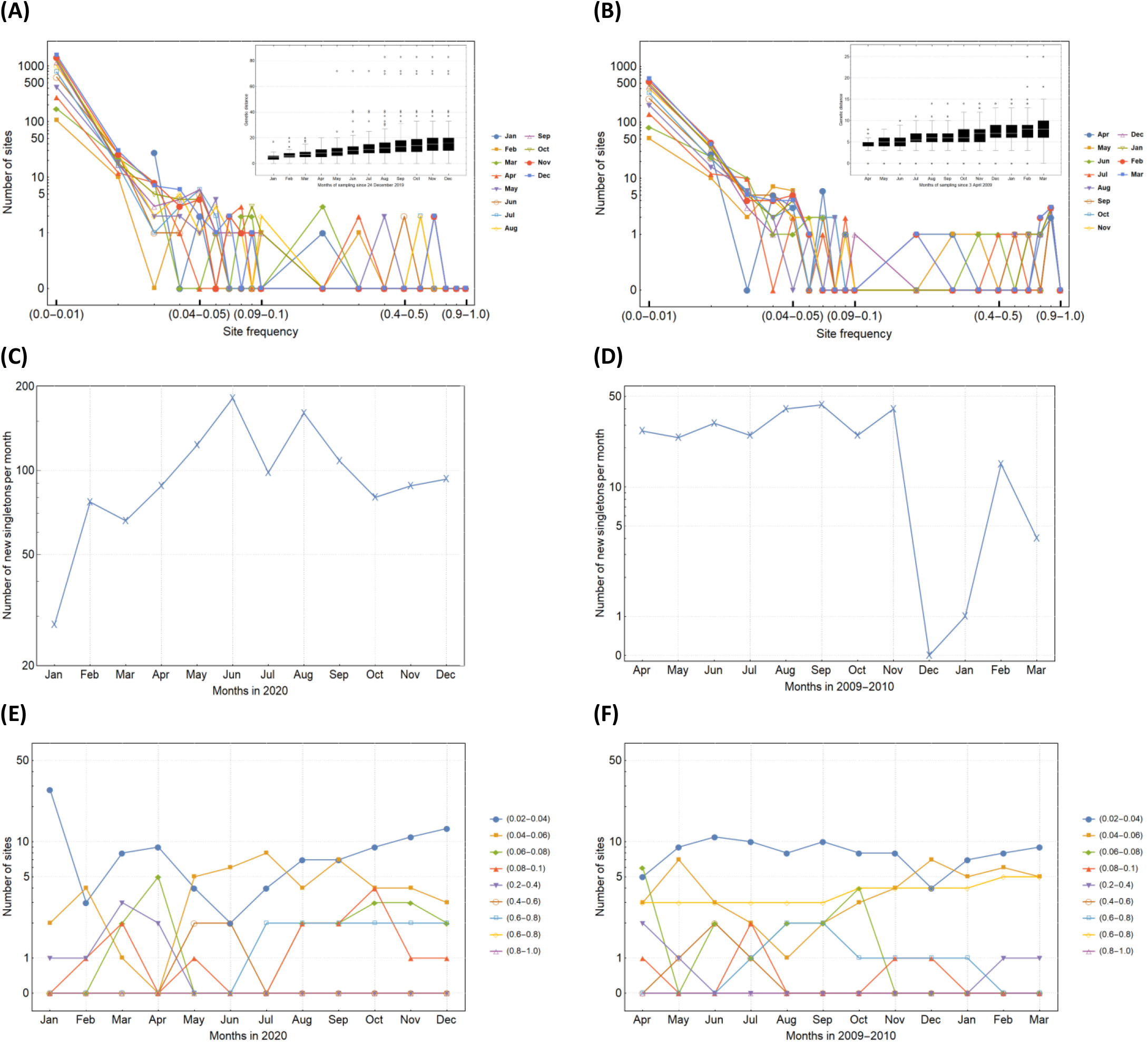
**(A, B)** Site frequency spectrum and genetic distance (in the inset) over time for SARS-CoV-2 (left column) and pH1N1 (right column). By comparing the main and ancestral alignments, each site in the main alignment is defined as invariant if it is identical to the ancestral nucleotide, or variant if it is not. Number of sites are counted based on the number of differences between the ancestral and main nucleotide positions. **(C, D)** Number of new single-nucleotide differences (singletons) added to the dataset per month. **(E, F)** Number of polymorphic sites in each frequency range over time.

